# Clinical efficacy of external beam radiotherapy complementing incomplete transarterial chemoembolization for hepatocellular carcinoma

**DOI:** 10.1101/2020.09.20.20197285

**Authors:** Sunmin Park, Won Sup Yoon, Mi Hee Jang, Chai Hong Rim

**Author notes:** Corresponding author: Chai Hong Rim, MD, Department of Radiation Oncology, Ansan Hospital, Korea University, 123 Jeokgeum-ro, Danwon-gu, Ansan, Gyeonggi-do, 15355, Republic of Korea. Biographical note: Sunmin Park is a radiation oncology specialist and a clinical instructor at the Korea University Medical College, who performed data curation and major drafting of this study. Won Sup Yoon is a head of the Radiation Oncology Department at Korea University Ansan Hospital. He published a number of studies regarding radiotherapy for breast cancer, liver cancer, and also dosimetry of radiotherapy. He supervised the manuscript and advised with his expertise and clinical experiences. Mi Hee Jang is a research nurse who performed main data recruitment and curation of the present study. Chai Hong Rim is a radiation oncologist and a passionate researcher who has published more than 30 articles as a primary or corresponding author in recent 3 years, including 11 articles about radiotherapy for hepatocellular carcinoma. He has also academic expertise in treating lung cancer, meta-analysis, and prevention oncology.

## Abstract

**Purpose:** External beam radiotherapy (EBRT) has been commonly applied as salvage or a combination locoregional modality after transarterial chemoembolization (TACE) for hepatocellular carcinomas (HCCs). This study reports oncologic outcomes and feasibility after application of the two modalities in our center.

**Methods:** Forty consecutive patients who underwent EBRT due to incomplete responses of TACE were evaluated. Fourteen patients (35.0%) received stereotactic body radiotherapy (SBRT) and the remaining patients received conventionally fractionated radiotherapy (RT). A majority of patients who underwent SBRT received doses of 27 to 48 Gy in 3–4 fractions (median *EQD_2_: 57.0 Gy). Conventionally fractionated RT was performed with a median EQD_2_ of 47.8 Gy.

**Results:** The median follow-up duration was 14.4 months (range: 2.6–83.0 months). A majority (77.5%) of patients were regarded as having Child-Pugh grade A. The median tumor size was 3.4 cm (range: 0.8–20.1 cm). Ten patients (25.0%) had thrombosis at a main portal branch. The 1- and 2-year overall survival (OS) and progression-free survival (PFS) rates were 82.2% and 42.1% and 55.8% and 32.1%, respectively. The local control rates were 89.1% and 89.1% at 1 and 2 years, respectively. The albumin level was a significant factor affecting OS (*p* = 0.002), and the BCLC stage significantly affected PFS (*p* = 0.001). Intrahepatic, out-of-field recurrence was the main cause of disease progression (60.0%), and distant metastasis developed in 12 patients (30.0%) during follow-up. Non-classic radiation-induced liver disease was seen in five (12.5%) patients, and two (5%) patients experienced grade ≥ 3 hepatic toxicities.

**Conclusions:** EBRT after incomplete TACE was feasible and yielded favorable oncologic outcomes. However, disease progression related to intrahepatic failure remained a hindrance.

*EQD2: Equivalent dose in 2 Gy per fraction radiotherapy

## Introduction

Transarterial chemoembolization (TACE) is clinically the most commonly applied locoregional treatment for hepatocellular carcinoma (HCC) (Park et al. 2015; Raoul et al. 2011). Given the established overall survival (OS) gain for patients with unresectable HCCs (Llovet et al. 2002; Lo et al. 2002), TACE has been solely recommended for intermediate stage cases in the Barcelona Clinic of Liver Cancer system (EASL 2018) and has been suggested as a primary locoregional modality for inoperable HCCs in many international guidelines (KLCA 2019; NCCN. 2020; S. Park et al. 2020).

The limitation of TACE is that complete remission and sustained local control is difficult to achieve, except for early HCCs such as single and small tumors (≤ 3 cm) (Henry et al. 2013; J. H. Shim et al. 2010). In a recent meta-analysis by Lencioni et al (Lencioni et al. 2016), progression-free survival (PFS) after TACE for HCCs was as mediocre as 24% at 2 years, and the majority of lesions progressed in a year.

External beam radiation therapy (EBRT) has been increasingly applied for HCC, as CT-based planning has enabled selective tumor irradiation while sparing normal liver (Rim and Yoon 2018). EBRT is advantageously performed regardless of tumor location and can be applied to tumors with locations where RFA is difficult to apply (e.g., tumors near major vessels or the diaphragm) (J. Lee et al. 2020; Rim et al. 2019). While standard salvage or complementing options after incomplete TACE have yet to be established, EBRT has been applied for these purposes in real-world experiences (Choi et al. 2014; Kang et al. 2012; Kim et al. 2006; Oh et al. 2010; S. J. Shim et al. 2005; Zhong et al. 2014).

This study aimed to report the clinical experience in our center, a middle-sized tertiary hospital in South Korea, in performing EBRT for HCCs to complement or salvage incomplete TACE. Related literature and suggestions for future treatment and research have also been discussed.

## Methods

### Patients recruitment and evaluation

A chart review was performed for HCC patients who underwent external radiation therapy (RT) between March 2010 and May 2019 (IRB number: 2020AS0076). Inclusion criteria were: 1) an initial diagnosis of primary HCC or recurrence; 2) an incomplete response after TACE (e.g., remnant viable lesion present or inadequate lipiodol uptake in the tumor; 3) age 18 years; 4) Child-Pugh (CP) class A or B; 5) ≥ 25 Gy of radiation dose; and 6) no evidence of uncontrolled lesions at any other sites than primary liver. Oncologic outcomes including survival outcomes, tumor response, and local control were evaluated. Tumor response was assessed using the Modified Response Evaluation Criteria in Solid Tumors (mRECIST) (Lencioni and Llovet 2010). We included patients who had undergone conventional fractionated RT and stereotactic body radiation therapy (SBRT). Complications related to RT were evaluated according to the Common Terminology Criteria for Adverse Events (CTCAE; version 4.03). Non-classic radiation-induced liver disease (RILD) was defined as elevated liver transaminase levels more than five times the upper normal limit or CP score worsening by ≥ 2 within 3 months after RT. Local control was defined as no new lesion or no increase in the tumor size in the treated area (in-field failure). Intrahepatic recurrence was defined as the development of a new lesion outside the treated area, but within the liver (out-of-field failure). Disease progression was defined as the development of intrahepatic recurrence and/or distant recurrence at any site outside of the liver. Overall survival, PFS, and recurrence-free survival (RFS) were estimated from the start date of RT to the date of death, the last follow-up examination, or to the date of tumor progression and recurrence, respectively. All patients had follow-ups at 3- or 6-months intervals after treatment. At each follow-up, a detailed questionnaire on clinical status and physical examination were administered and liver magnetic resonance imaging (MRI) or dynamic computed tomography (LDCT) were performed.

### Statistics

The Kaplan–Meier method was used to evaluate the probability of cumulative survival. Univariate and multivariate Cox proportional hazards models were used to evaluate the association of variables with survival outcomes and local control. The backward elimination method was used to select the principal factors in multivariate analyses. Univariate logistic regression analysis was performed to compare clinical parameters for non-classic RILD. Statistical Package for the Social Sciences version 21.0 were used for all statistical analyses (IBM Corp., Armonk, NY, USA).

### Radiotherapy procedure

CT simulation was performed using a Philips Brilliance Big Bore CT (Philips Healthcare, Cleveland, OH, USA). A compression belt was applied at the mid-abdomen of the patients who were lying on the Body Pro-Lok™ (Body Pro-Lok, CIVCO, USA) in a supine position. The patients were immobilized using Vac-L ok (KIKWANG MEDICAL, KOREA) and wing board (CIVCO, USA). Four-dimensional CT (4DCT) was scanned to a slice thickness of 2–3 mm using the Varian RPM system (Varian Medical Systems, Palo Alto, CA) to monitor and record a respiratory signal, specifically the rise and fall of the anterior abdominal surface. A 4DCT image was taken while abdominal compression was performed. After scanning, the 4DCT image data sets were sorted into 10 phases such that the 0% respiratory phase corresponds to peak inhalation and the 50% phase to peak exhalation. A total of 10 phases are evenly distributed throughout the respiratory cycle. The GTV can consist of the entire HCC or part of the HCC and vascular invasion at the discretion of the investigator. The GTV was contoured in the CT equating to the 50% phase, and the internal target volume (ITV) was set to be expanded by reflecting the movement in all phases.

Planning target volume (PTV) was set with a margin of 5–7 mm from ITV. Dose constraints to organs at risk (OAR), including the duodenum, liver, spinal cord, and esophagus, were adopted according to the Quantitative Analyses of Normal Tissue Effects in the Clinic (QUANTEC) (Bentzen et al. 2010). In case of SBRT, a dose was prescribed to an isodose covering at least 95% of the PTV. If OAR constraints are not met, then the 95% isodose was relaxed or the total dose was reduced according to the clinicians’ discretion. The dose maximum within the PTV was preferably were between 110% and 140% of the prescribed dose. For IMRT, the target dose was prescribed to the PTV more than 95% of the prescribed dose to at least 95% of the PTV. For 3DCRT planning, 3–5 portal beams with planar or non-coplanar arrangement were used. Dosimetric margins around the PTV were selected so that the 80% isodose line encompasses 100% of the PTV. In addition, PTV received 95%–107% of the prescribed total dose. The biologically effective dose (BED) and equivalent dose in 2 Gy per fraction radiotherapy (EQD2) were calculated from the prescription dose using α/β ratio of 10. Gating during treatment was not performed because ITV reflected movements of all phases in free breathing. The setup was performed using a tattoo applied to the patient, and daily pre-treatment cone-beam computed tomography (CBCT) matching was verified by a physician. Image guidance was performed by matching the liver contour and lipiodolized GTV as fiducial. If matching was satisfactory, then treatment was performed after applying the couch shift.

## Results

### Patient characteristics

Between March 2010 and May 2019, 44 patients were treated with RT due to incomplete response of TACE. Of them, 40 patients (46 lesions) met all the enrollment criteria and were included in this analysis. Most patients were males (82.5%). Median age was 60 years (range, 43.0–77.0 years) at the initiation of RT. The longest diameter of the tumors was 0.8–20.1 cm, with median value of 3.42 cm. The tumor volume was 4.0–1939.4 cm^3^, with median value of 94.1 cm^3^. A majority (77.5%) of patients were regarded as having CP class A, and the remaining were regarded as having CP class B. Ten patients (25.0%) had thrombosis at a main portal branch and two patients (5.0%) in a segmental branch. Patients had undergone conventionally fractionated RT with doses of 25–65 Gy (median EQD2: 47.8 Gy) in 8–27 fractions. SBRT was performed with doses of 27–48 Gy (median EQD2: 57.0 Gy) in 3–4 fractions (Table 1). Thirteen of 40 patients (32.5%) received systemic chemotherapy (sorafenib and regorafenib) during the entire follow-up period. None of the 13 patients received systemic therapies simultaneously with RT; all received these after completion of RT.

**Table 1.** Patient characteristics

| Variables | No. of patients (%) |
| --- | --- |
|  | (n = 40) |
| Sex |  |
| Male | 33 (82.5%) |
| Female | 7 (17.5%) |
| Age (years) |  |
| Median (range) | 60 (43–77) |
| BCLC |  |
| A | 9 (22.5%) |
| B | 12 (30.0%) |
| C | 19 (47.5%) |
| ECOG performance status |  |
| 0 | 13 (32.5%) |
| 1 | 26 (65.0) |
| 2 | 1 (2.5%) |
| Child-Pugh Class |  |
| A | 31 (77.5%) |
| B | 9 (22.5%) |
| Viral Etiology |  |
| Hepatitis B virus | 25 (62.5%) |
| Hepatitis C virus | 6 (15.0%) |
| Non B, Non C | 9 (22.5%) |
| Alpha-fetoprotein (ng/mL) |  |
| Range | 2–78209 |
| ≤200 | 29 (72.5%) |
| >200 | 11 (29.3%) |
| Portal vein thrombosis |  |
| Yes, main branch | 10 (25.0%) |
| Yes, segmental branch | 2 (5%) |
| No | 28 (70%) |
| Number targets for RT |  |
| Single lesion | 37 (90.2%) |
| Two lesions | 3 (7.3%) |
| Four lesions | 1 (2.4%) |
| Tumor size (cm)* |  |
| Median (range) | 3.4 (0.8–20.1) |
| 0.7–3.0 | 20 (43.5%) |
| 3.1–5.0 | 9 (19.6%) |
| 5.1–10.0 | 11 (23.9%) |
| 10.1–25.0 | 6 (13.0%) |
| Tumor volume (cm<sup>3</sup>) * |  |
| Median (range) | 94.1 (4.0–1939.4) |
| Prior treatments |  |
| TACE only | 18 |
| TACE, RFA | 12 |
| TACE, PEI | 1 |
| TACE, RFA, PEI | 1 |
| TACE, RT | 1 |
| Resection, TACE | 4 |
| Resection, TACE, RFA | 2 |
| HAIC, RFA | 1 |
| HAIC, TACE, PEI | 1 |
| RT regimen * |  |
| Conventional fractionated RT | 26 (65.0 %) |
| 3DCRT | 14 (35.0%) |
| IMRT | 12 (30.0%) |
| SBRT | 14 (35.0%) |
| RT dose, median (range) * |  |
| EQD2 | 48.6 (26.0–89.4) |
| BED (Gy<sub>10</sub>) | 58.3 (31.3–107.3) |
| Treatment aim * |  |
| Palliative | 5 (10.9%) |
| Definitive | 35 (87.5%) |

\* Assessed for each lesion
BCLC, Barcelona Clinic of Liver Cancer; ECOG, Eastern Cooperative Oncology Group; RT, radiotherapy; TACE, transarterial chemoembolization; HAIC, hepatic arterial infusion chemotherapy; RFA, radiofrequency ablation; PEI, percutaneous ethanol injection; 3DCRT, 3-dimensional conformal radiotherapy; IMRT, intensity modulated radiotherapy; SBRT, stereotactic body radiotherapy; EQD2, equivalent dose in 2Gy per fraction; BED, biologically effective dose.

### Survival analysis

The median follow-up duration was 14.4 months (range, 2.6–83.0 months) for all patients, and 23 of the 40 patients survived during the follow-up period. The median OS was 15.0 months (range, 2.6–100.6 months). The 1- and 2-year OS rates were 82.2% and 55.8%, respectively (Figure 1). The 1- and 2-year PFS rates were 42.1% and 32.1% (Figure 1), respectively. For OS, the albumin level was the sole significant factor in both univariate and multivariate analyses (*p* = 0.002 in multivariate analysis, Table 2). The BCLC stage was the only significant prognostic factor (*p* = 0.006 in multivariate analysis) for PFS in both univariate and multivariate analysis (Table 3).

**Table 2.**
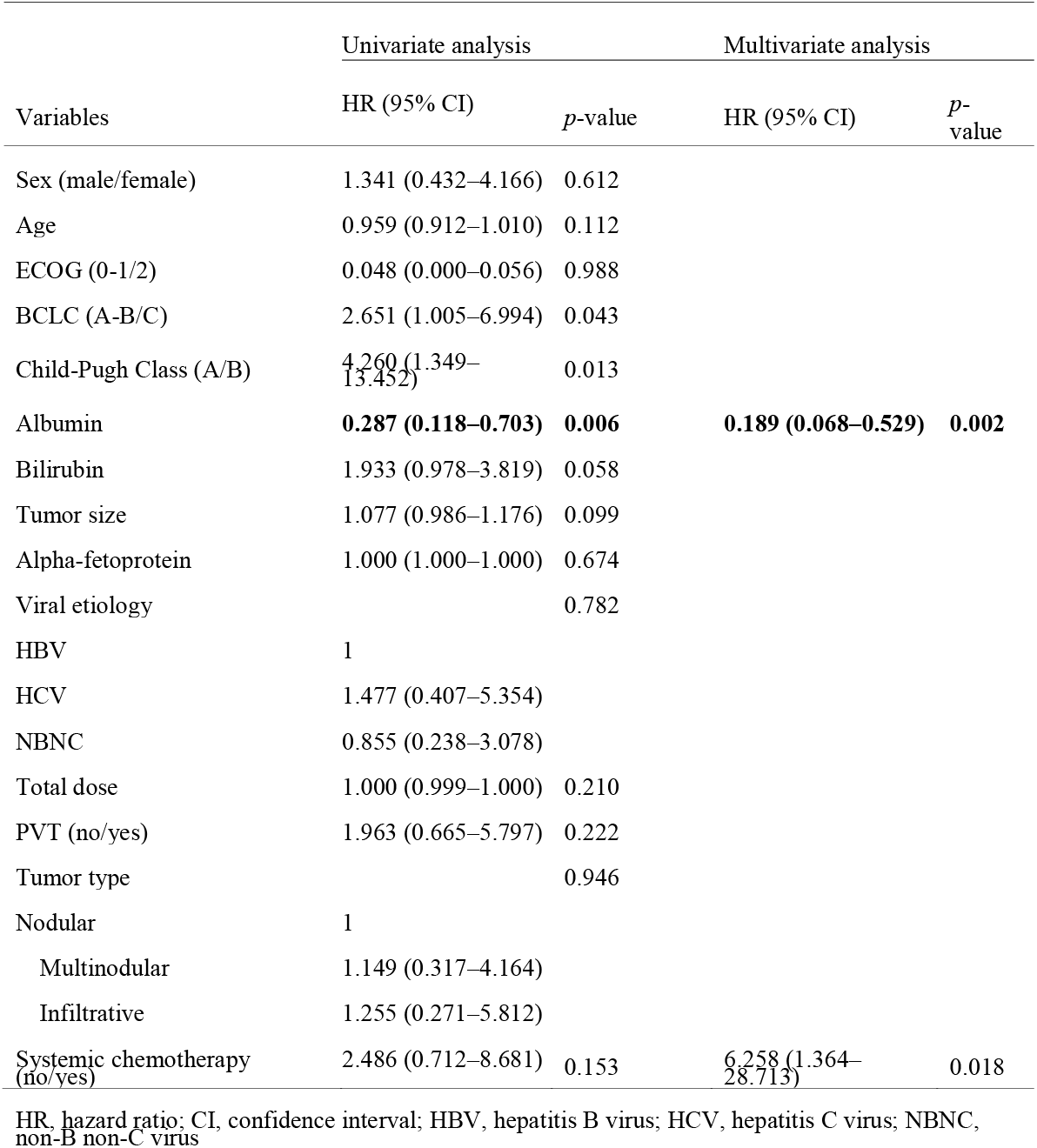
Predictive factors influencing overall survival

**Table 3.** Predictive factors influencing progression free survival

| Variables | Univariate analysis |  | Multivariate analysis |  |
| --- | --- | --- | --- | --- |
|  | HR (95% CI) | *p*-value | HR (95% CI) | *p*-value |
| Sex (male/female) | 0.311 (0.073–1.328) | 0.115 |  |  |
| Age | 0.980 (0.942–1.020) | 0.980 |  |  |
| ECOG (0-1/2) | 2.703 (0.351–20.810) | 0.602 |  |  |
| BCLC (A-B/C) | **4.121 (1.729–9.823)** | **0.006** | **3.530 (1.070–11.642)** | **0.001** |
| Child-Pugh Class (A/B) | 1.167 (0.465–2.933) | 0.742 |  |  |
| Albumin | 1.327 (0.640–2.751) | 0.448 | 2.637 (0.962–7.228) | 0.059 |
| Bilirubin | 1.549 (0.874–2.745) | 0.134 | 2.639 (1.165–5.978) | 0.020 |
| Tumor size | 1.046 (0.971–1.127) | 0.238 |  |  |
| Alpha-fetoprotein | 1.000 (1.000–1.000) | 0.112 | 1.000 (1.000–1.000) | 0.019 |
| Viral etiology |  | 0.352 |  | 0.098 |
| HBV | 1 |  | 1 |  |
| HCV | 1.763 (0.590–5.272) |  | 2.746 (0.830–9.080) |  |
| NBNC | 0.633 (0.214–1.874) |  | 0.469 (0.148–1.489) |  |
| Total dose | 1.000 (0.999–1.000) | 0.185 |  |  |
| PVT (no/yes) | 2.535 (1.057–6.082) | 0.037 |  |  |
| Tumor type |  | 0.087 |  |  |
| Nodular | 1 |  |  |  |
| Multinodular | 2.805 (1.065–7.387) |  |  |  |
| infiltrative | 2.040 (0.765–5.442) |  |  |  |
| Systemic chemotherapy (no/yes) | 2.476 (1.102–5.564) | 0.023 |  |  |

HR, hazard ratio; CI, confidence interval; HBV, hepatitis B virus; HCV, hepatitis C virus; NBNC, non-B non-C virus

**Figure 1.**
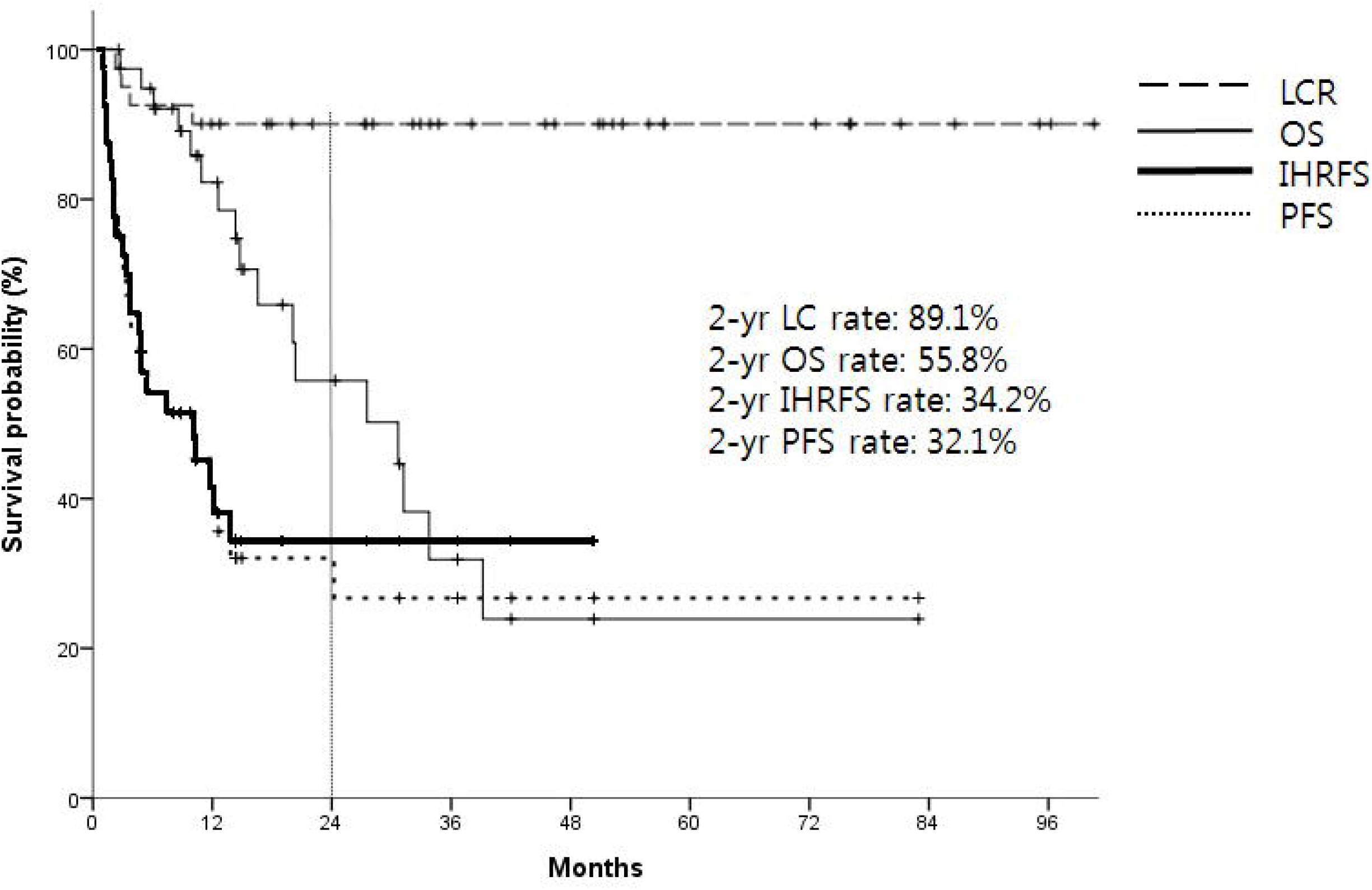
Kaplan-Meier Graph of overall survival (OS, progression free survival (PFS), local control rate (LCR), and intra-hepatic recurrence free survival (IHRFS)

### Tumor response and local control rate

In-field (target lesion) complete remission (CR) and partial response (PR) rates were 37.0% and 41.3%, respectively. Four patients (8.7%) were PD and six (13.0%) were SD. Overall in-field response (CR and PR) rate was 78.3%. The 1- and 2-year local control (LC) rates were 89.1% and 89.1%, respectively (Figure 1). For LC, the tumor type (nodular/multinodular/infiltrative) was a statistically significant factor in the univariate analysis (*p* = 0.02) and had borderline significance in the multivariate analysis (*p* = 0.077). The 1- and 2-year intrahepatic recurrence free survival (IHRFS) rates were 41.5% and 34.2%, respectively. The univariate and multivariate analyses revealed that the BCLC stage was the only significant factor for IHRFS (*p* = 0.003, and *p* = 0.003, respectively. Figure 1).

### Patterns of failure

Overall, 26 patients (65.0%) had treatment failures. Local failure (in-field failure) occurred in 5 of 46 lesions in four patients (10.0%). Intrahepatic out of-field failure was observed in 24 patients (60.0%), and distant failure (extrahepatic metastasis) occurred in 12 patients (30.0%) (Figure 2). The most common site for distant failure was the lungs, followed by the lymph nodes and bones.

**Figure 2.**
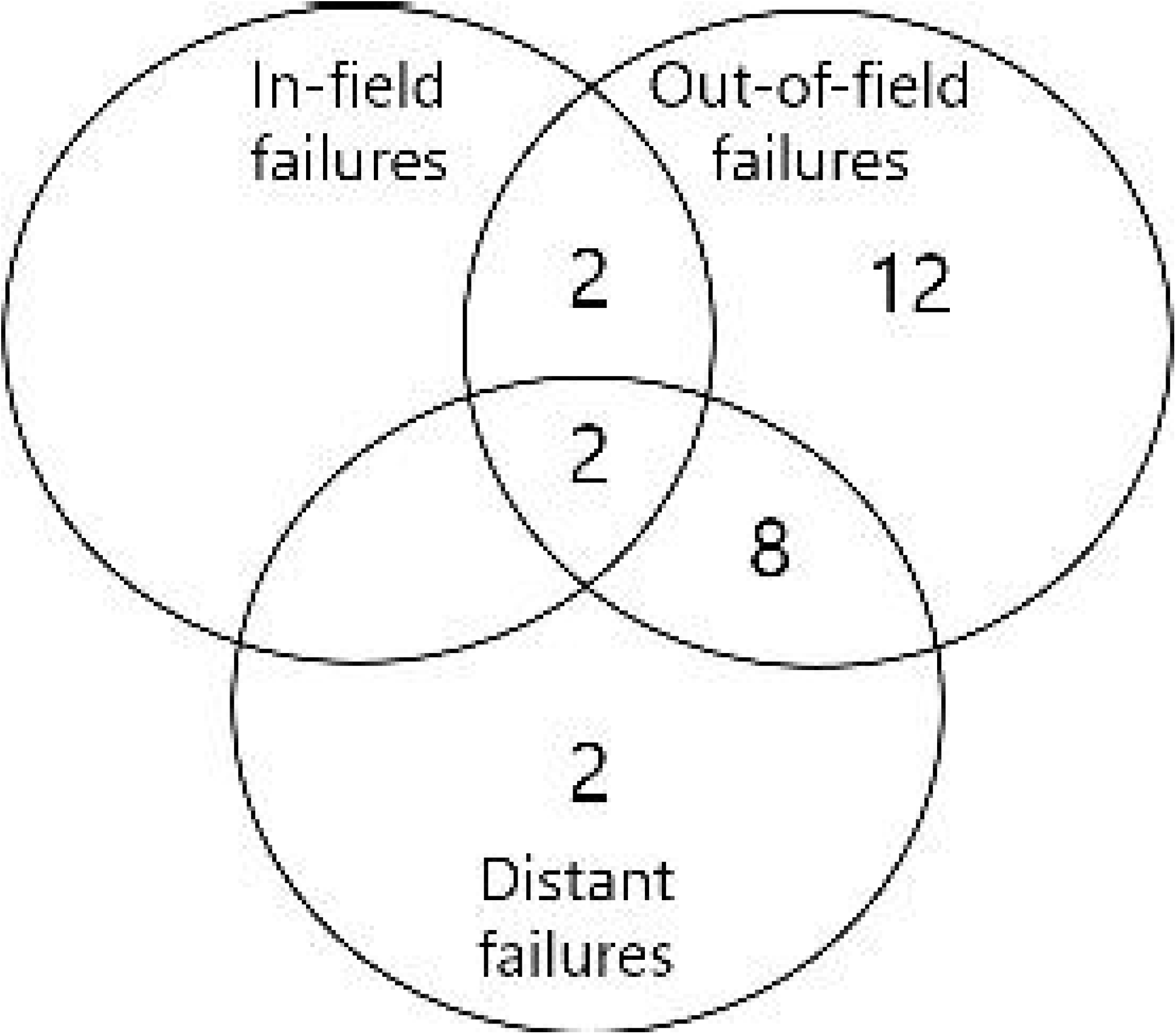
Patterns of failure after radiation therapy

### Treatment-related toxicities

All patients received planned RT without any interruptions due to intolerable side effects. No classic RILD was observed. Non-classic RILD was seen in five (12.5%) patients. Elevation of liver transaminase levels by more than five times was observed in two (5.0%) patients within 3 months after RT (Grade ≥ 3 hepatic toxicities; grade 3: one patient, grade 4: one patient). Two patients who experienced grade ≥ 3 hepatic toxicities improved spontaneously with conservative treatment. Three (7.5%) patients showed elevation in the CP score of ≥ 2. Of the five patients who experienced non-classic RILD, two underwent SBRT, two underwent 3DCRT, and one underwent intensity modulated radiotherapy (IMRT). In these patients, the median mean liver dose (MLD) was 13.3 Gy (range, 7.9–29.8 Gy) and the CP score was 6 (5–8) points. In the univariate analysis, none of the major clinical parameters were associated with non-classic RILD (Supplementary Table 1). Two patients experienced grade 3 thrombocytopenia (Table 4). One patient experienced diminution of platelet count to 20,000/ul after RT due to esophageal variceal bleeding, but this normalized after platelet concentrate transfusion and esophageal variceal ligation. The other patient, however, developed HCC rupture 2 months after RT and 1 month after consequent TACE, showing grade 3 thrombocytopenia with hemoperitoneum. After conservative care including transfusion, thrombocytopenia stabilized, but 3 weeks after HCC rupture, the patient died of asphyxia after vomiting. Severe gastrointestinal complications such as bleeding or perforation were not reported during the follow-up period.

**Table 4.** Toxicities after RT

|  | No. of patients (%) |  |  |  |  |
| --- | --- | --- | --- | --- | --- |
|  | Grade 1 | Grade 2 | Grade 3 | Grade 4 | Grade 5 |
| Fatigue | 4 (10.0%) |  |  |  | - |
| Anorexia | 2 (5.0%) | - | - | - | - |
| Nausea/vomiting | 4 (10.0%) |  | - | - | - |
| Abdominal pain | 9 (22.5%) | 3 (7.5%) | - | - | - |
| Diarrhea | 1 (0.4%) | - | - | - | - |
| Hyperbilirubinemia | 9 (22.5%) | 7 (17.5%) | - | - | - |
| Thrombocytopenia | 10 (25.0%) | 11 (27.5%) | 2 (5.0%) | - | - |

## Discussion

The study results demonstrate that application of EBRT after incomplete TACE can confer a favorable response and sustained in-field tumor control. A majority of complications were transient and mild, and serious toxicity (e.g., grade ≥ 3) was rare. Prognostic factors, including the albumin level and BCLC stage affecting the OS and PFS, were correlated with known HCC biology and previous studies (Klein and Dawson 2013); the albumin level can reflect not only the nutritional status but also the liver function and is an independent prognostic factor of oncologic outcomes (Bagirsakçi et al. 2017; Liu et al. 2017; Wu et al. 2019).

Considering the result of our study and the previous literature, in-field local control was mostly favorable after incomplete TACE (Table 5) (Chiang et al. 2019; Chihwan. Choi et al. 2014; Jacob et al. 2015; Kang et al. 2012; Kibe et al. 2020; Kim et al. 2006; Oh et al. 2010; S. J. Shim et al. 2005; Zhong et al. 2014). Although the study by Choi et al (Chihwan. Choi et al. 2014). Reported a relatively high in-field failure rate, high incidences of infiltrative or multinodular tumor type (80%) and BCLC C patients (45%) might have affected the result. Since the subject of literature is a salvage setting, many of the referenced studies involved a portion of patients with relatively large tumor or portal vein thrombosis. A majority of studies prescribed a dose of around 40–60 Gy in EQD2_10Gy_ in both SBRT and conventional schemes, indicating a difficulty in aggressive dose escalation. In the studies by Jacob et al. (Jacob et al. 2015) and Kang et al. (Kang et al. 2012), high-dose SBRT achieved good local control. However, considering that locally advanced cases are commonly involved and that HCC has a relatively high α/β ratio, which is benefited by the fractionated scheme (∼15Gy) (Tai et al. 2008), conventionally or moderately fractionated RT could be also suitable options reflected by favorable in-field control in studies including ours.

**Table 5.**
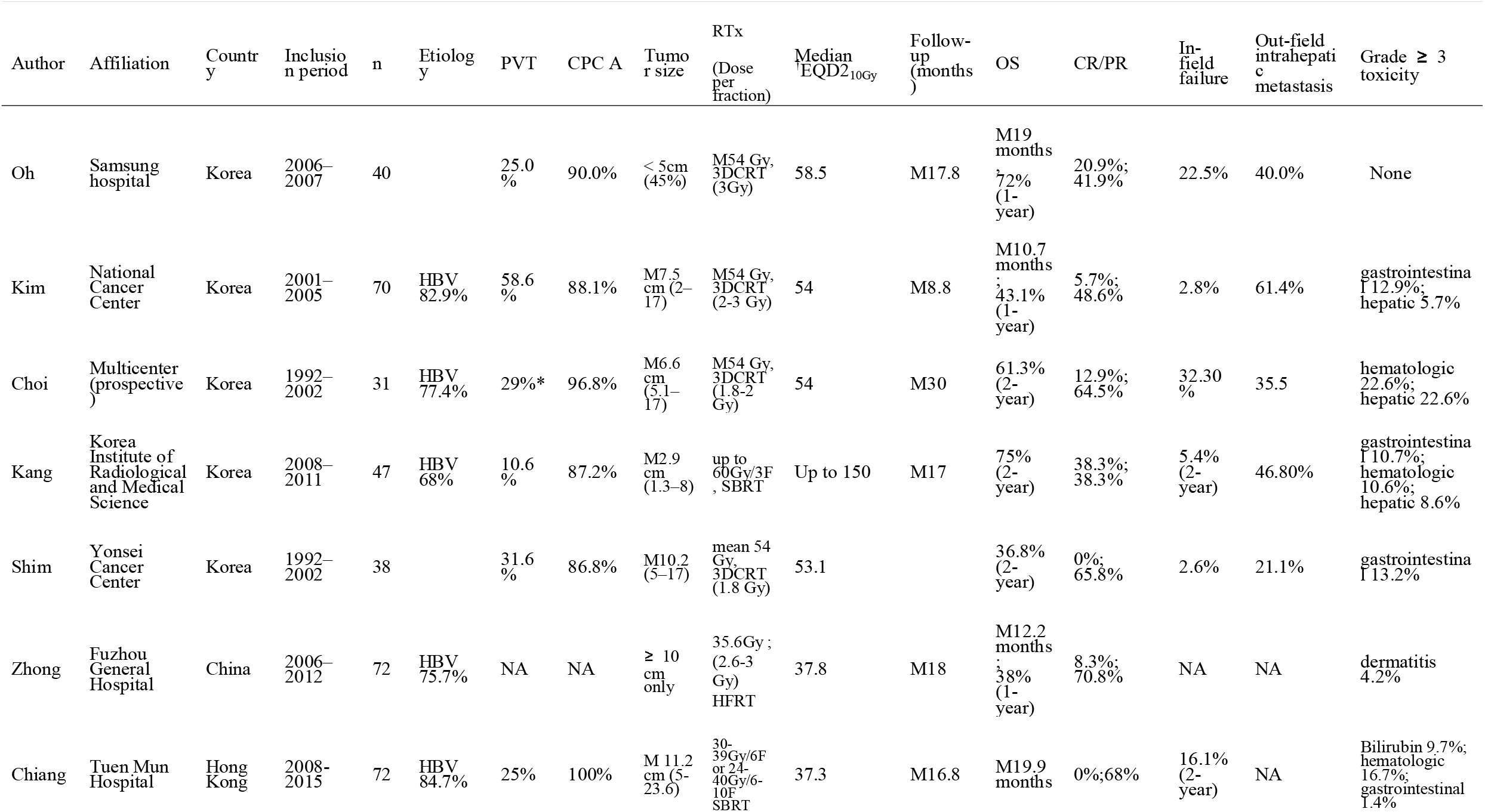
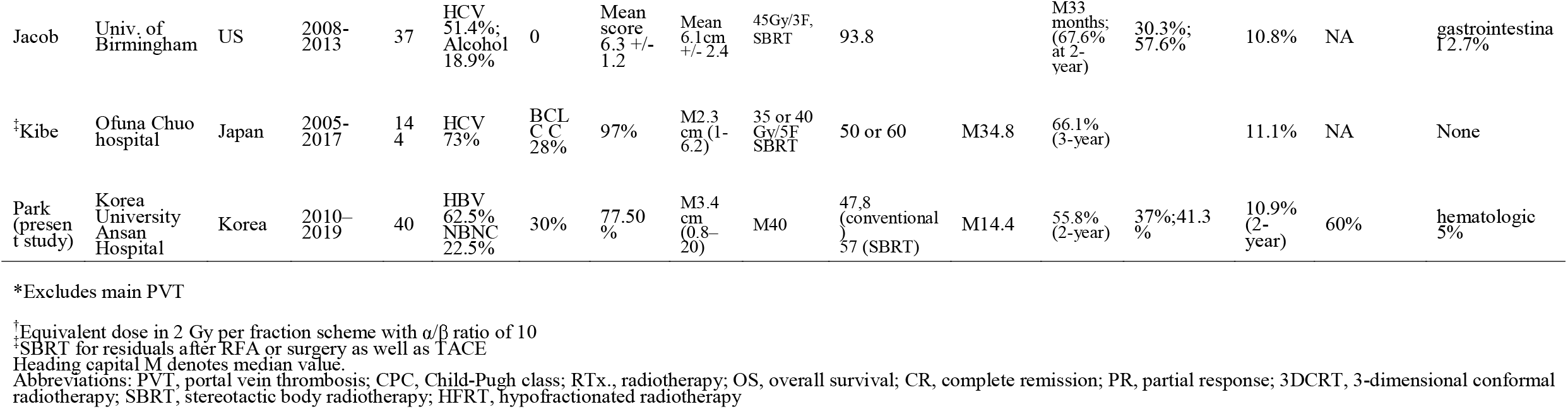
Selected previously published studies with correlating subjects

Out-of-field intrahepatic metastases were burdensome in most of the studies, including the present one and approximately ranged from 405 to 60%. Such a result sheds light on the role of systemic treatment; however, literature regarding the concomitant use of systemic treatment and EBRT for HCC is still scarce. In a phase 2 study of combined sorafenib and EBRT for HCC, oncologic efficacy was moderately favorable (2-year OS: 32%, response rate: 55%), but intrahepatic out-of-field progression was still problematic (30%) (Chen et al. 2014). In a comparative study by Wada et al. (Wada et al. 2018), combined use of sorafenib and EBRT showed a comparable toxicity rate to sorafenib alone, but the 2-year PFS was <40%.

Intrahepatic progression has been deemed one of the most difficult challenges regarding HCC treatment. Additional studies are still necessary, but there are several promising scenarios. Preclinical evidence of synergistic effects whereby EBRT affects tumor microenvironments, enhancing the effects of immune checkpoint inhibitors, has emerged (Choi et al. 2019). Recently, the combined application of anti-VEGF and an immune checkpoint inhibitor demonstrated significantly superior oncologic outcomes to sorafenib (Finn et al. 2020). Nivolumab also showed promising results in its phase 1/2 trials for advanced HCCs; the response rate was 20% and the median duration of response was 9.9 months (Melero et al. 2017). Several trials of combined treatment of EBRT and immune checkpoint inhibitors (NCT0382102, NCT03203304, NCT0331672) are ongoing, from which contributions to reduce intrahepatic progression are expected. In addition, surgical conversion after EBRT combined with arterial directed therapy might yield excellent outcomes. In a series reported from a Korean tertiary center, although the conversion rate to surgery was <20%, the median OS of patients who underwent conversion surgery after EBRT exceeded 5 years (Byun et al. 2019; H. S. Lee et al. 2014). Therefore, consequent surgical resection, which might prevent further hepatic progression, might be considered for favorable responders after EBRT.

In addition to perspectives of numerical oncologic outcomes, EBRT is a practical and feasible option for locally advanced HCCs for which other locoregional modalities are difficult to apply for anatomical reasons. We selected two cases showing significant improvements initially referred as intractable cases. The case depicted in Figure 3 upper showed complete remission after EBRT for infiltrative HCC involving a major vessel and the middle portion between the bilateral lobes, which is deemed very difficult for the application of any locoregional modalities. The case in Figure 3 lower shows an impressive response to EBRT for a huge tumor for which previous TACE was not very efficacious and that subsequent TACE had greater effects inside the tumor (hypovasculated with necrosis) partly due to vasculature remodeling after EBRT (Castle and Kirsch 2019; El Kaffas et al. 2013).

**Figure 3.**
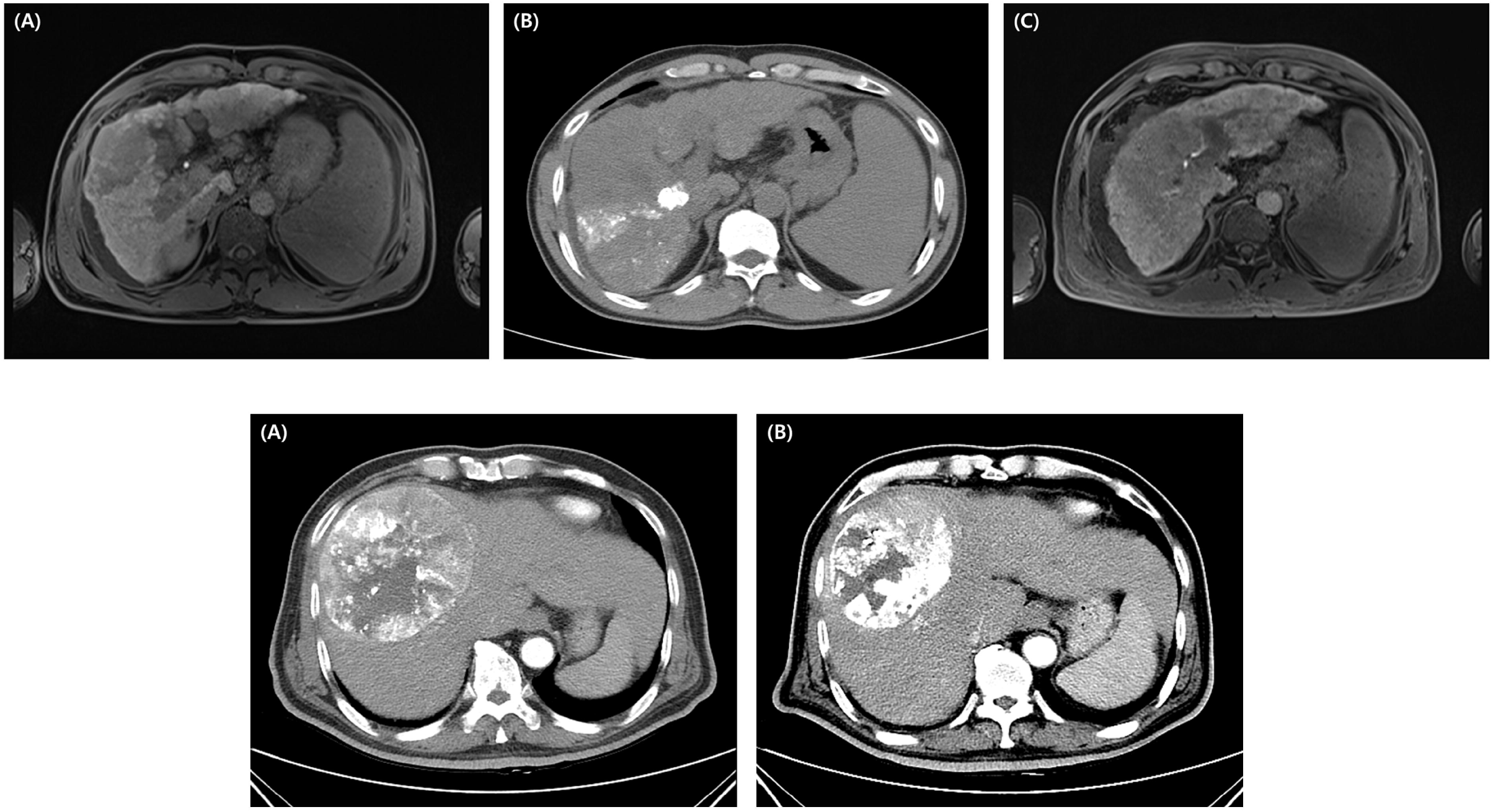
Case 1 (patient no. 1, upper figures) (A) This case involved a patient in her 40s, with HCC with hepatitis B and related cirrhosis. Liver magnetic resonance image (MRI) showed a 9.2 cm-sized infiltrative tumor with PVT. (B) First, TACE was performed after diagnosis. Lipodol was compactly tagged in the right PVT, but incomplete TACE was performed on the remaining lesions. He underwent radiotherapy with 54 Gy in 27 fractions. (C) Follow-up imaging at 3 months after radiation therapy. The patient has achieved CR without any toxicity. Case 2 (patient no. 2, lower figures) (A) A main in his 50s, was diagnosed with hepatitis-B-related HCC. Liver dynamic CT showed a 13.4 cm-sized huge viable HCC in hepatic segment 8. After two TACE treatments, there remained a persistent tumor in segment 8. He underwent radiotherapy with 40 Gy in 16 fractions. (B) At follow-up imaging at 3 months, the longest diameter was reduced to 10.4 cm, and consequent TACE affected more of the inside of the large tumor after radiotherapy.

Our study has some limitations, including its retrospective design and the small number of patients and clinical heterogeneity. These limitations are shared by several other studies with correlated subjects. The number of patients included in future studies should be increased through means such as multi-center recruitment, consequently enabling more specific studies with less heterogeneous subgroups. As mentioned earlier, studies regarding a combined treatment of EBRT and immune checkpoint inhibitors and consequent resection for favorable responders are strongly warranted.

## Conclusion

EBRT after incomplete TACE is a feasible option yielding favorable tumor response and sustained local control. However, intrahepatic progression remains a hindrance conferring a mediocre level of survival. Future studies should be undertaken to prevent intrahepatic progression, investigating combined treatment with novel systemic treatment and consequent surgical resection of favorable responders of EBRT.

## Data Availability

The data that support the findings of this study are available on request from the corresponding author.

## Acknowledgement

This study is supported by National Research Fund of Korea (NRF-2019M2D2A1A01031560).

## Ethical approval and informed consent

Our study is evaluated and approved by the Institutional Review Board of Korea University Medical Center (IRB number: 2020AS0076), and informed consent was waived because our study did not pose any harm to the patients involved and no personally identifiable information was used. References: see the journal’s instructions for authors for details on style

